# Efficient and Practical Sample Pooling for High-Throughput PCR Diagnosis of COVID-19

**DOI:** 10.1101/2020.04.06.20052159

**Authors:** Haran Shani-Narkiss, Omri David Gilday, Nadav Yayon, Itamar Daniel Landau

## Abstract

In the global effort to combat the COVID-19 pandemic, governments and public health agencies are striving to rapidly increase the volume and rate of diagnostic testing. The most common form of testing today employs Polymerase Chain Reaction in order to identify the presence of viral RNA in individual patient samples one by one. This process has become one of the most significant bottlenecks to increased testing, especially due to reported shortages in the chemical reagents needed in the PCR reaction.

Recent technical advances have enabled High-Throughput PCR, in which multiple samples are pooled into one tube. Such methods can be highly efficient, saving large amounts of time and reagents. However, their efficiency is highly dependent on the frequency of positive samples, which varies significantly across regions and even within regions as testing criterion and conditions change.

Here, we present two possible optimized pooling strategies for diagnostic SARS-CoV-2 testing on large scales, both addressing dynamic conditions. In the first, we employ a simple information-theoretic heuristic to derive a highly efficient re-pooling protocol: an estimate of the target frequency determines the initial pool size, and any subsequent pools found positive are re-pooled at half-size and tested again. In the range of very rare target (<0.05), this approach can reduce the number of necessary tests dramatically, for example, achieving a reduction by a factor of 50 for a target frequency of 0.001. The second method is a simpler approach of optimized one-time pooling followed by individual tests on positive pools. We show that this approach is just as efficient for moderate target-product frequencies (0.05<0.2), for example, achieving a two-fold in the number of when the frequency of positive samples is 0.07.

These strategies require little investment, and they offer a significant reduction in the amount of materials, equipment and time needed to test large numbers of samples.

We show that both these pooling strategies are roughly comparable to the absolute upper-bound efficiency given by Shannon’s source coding theorem. We compare our strategies to the naïve way of testing and to alternative matrix-pooling methods. Most importantly, we offer straightforward, practical pooling instructions for laboratories that perform large scale PCR assays to diagnose SARS-CoV-2 viral particles. These two pooling strategies may offer ways to alleviate the bottleneck currently preventing massive expansion of SARS-CoV-2 testing around the world.

## Introduction

In the global effort to fight the coronavirus pandemic, medical teams and laboratories presented with the task of diagnosis are currently encountering unprecedented numbers of samples, and simultaneously facing shortages of time, personnel, materials and laboratory equipment.

The need to scale up diagnostic assays for many thousands of patient samples has been addressed with cutting edge molecular tools such as RNA-Seq with multiplex barcoding^1^ and serological test might soon be able to test for the immune status of patients. However, these tools may not be readily available to implement in many places, and they often require higher expertise or are less accurate than regular PCR tests commonly used. A simpler way to scale up diagnostic assays can be found in the method of High-Throughput PCR via sample pooling, used in genetic research as a practical way to reduce the cost of large-scale studies^4^.

The most common current procedure for diagnosing the presence of SARS-CoV-2 begins with collection of a viral sample by a nasopharyngeal swab and/or an oropharyngeal swab from the patient. After lysis, the disintegration of cells/viral membrane within the sample, the detection procedure involves two stages:

1) RNA extraction which contains viral as well as Human RNA (later used for extraction control) are extracted using standard RNA extraction procedures^5^.

2) Single-step Reverse Transcription – quantitative Polymerase Chain Reaction (Single Step RT-qPCR) is performed on viral RNA and human control^5^. Briefly, in RT-qPCR reaction, the extracted RNA is first reverse transcribed to a double stranded cDNA template. Next, a reaction is repeated in cycles, amplifying the target cDNA fragment exponentially, by doubling the amount of that fragment in each cycle. In a typical qPCR reaction this amplification is repeated for ∼42 cycles. The cycle in which the fluorescent signal crosses a certain threshold is linked to the starting concentration of the target cDNA. In practice, if the presence of amplified viral cDNA is detected in a significant amount before a certain cycle number (e.g. 30), and given that the human cDNA (extraction control) was also detected in the sample, the patient is declared positive. If amplified viral cDNA is not detected or only detected in very late cycles (e.g. 40) then the patient is declared negative.

This RT-qPCR reaction is the most consuming stage of the process both in terms of time and reagents. Recent reports from around the world identify the RT-qPCR reaction as one of the primary bottlenecks in the entire COVID-19 testing enterprise – each sample tube tested requires chemical reagents that are increasingly in short supply as the number of PCR reactions performed globally grows tremendously^11,12^. Laboratories have begun to demonstrate that SARS-CoV-2 can be detected in RT-qPCR performed on pooled samples, despite potential dilution^2^. The input to pooling methods is RNA extracted from samples individually, although they may be also used to combine “raw” patient samples, even before lysis and extraction. Their aim is to identify the presence of viral RNA without the need to perform the RT-qPCR reaction on every sample individually.

To understand the advantages of a pooling approach, consider a laboratory receiving *N* = 1000 samples, with a frequency *p* = 1/1000 of positive cases. Using a naïve, one-by-one approach, 1000 tests will need to be performed. However, if the samples are pooled together, for example, into 10 different batches of *b* = 100 samples each, it is probable that 9 out of 10 batches will show a negative result. Each negative result, obtained by a single RT-qPCR reaction, determines that 100 individual samples are negative without the need for individual testing. Samples in batches that yielded a positive result may be either processed individually, or further divided to smaller batches. Both approaches will improve the testing efficiency significantly.

However, what happens when the frequency of positive samples (*p*) rises? It is clear that a strategy of pooling 100 samples together will not be beneficial for higher *p*, say 0.2. In this case, we can be certain that every batch will show positive, and therefore the chance that the pooling will yield any information at all is essentially zero. In practice the frequency of positive samples has varied greatly from country to country, depending on the criterion for testing and the stage of the pandemic at time of testing. For example, as of March 15^th^, there had been 167,009 tests performed in Germany, with 6540 positives, a rate of *p=*0.04^8^. On the other hand, as of March 31^st^, there had been 956,481 tests performed in the US, with 162,399 positives, a rate of *p*=0.*17*, more than four times higher^7^. These rates additionally vary across cities and regions, and also change over time. Expanding testing to surveys in asymptomatic populations is expected to significantly reduce the typical values of *p*. Thus, the practical efficiency of sample pooling requires developing laboratory protocols that adjust batch sizes for dynamically changing conditions.

Therefore, we set out to find a solution to the following problem: what is the optimal batch size for pooling samples, for any given *p*. We do this with two alternative approaches. The first approach calls for repeated pooling in several stages, and may be harder to implement in everyday lab work. However, it offers dramatic reductions in the number of tests required for very low values of *p*, and might be very useful for more advanced laboratories focusing on surveys of asymptomatic populations, where the appearance of positive examples is expected to be low. The second approach was in fact proposed as early as 1943^3^ and requires only one pooling step. Optimal initial batch size is calculated instantly, and then samples in all positive batches are to be further tested individually. This approach is very easy to implement, and as we will show, is very efficient for *p* values up to 0.2.

The problem we set to solve involves only 3 parameters; N is the number of samples available for the whole diagnostic assay, *p* represents the expected frequency of positive samples out of all samples (in practice, it should be calculated and updated on a daily basis, for each lab or for a given region/country); and *b*, the number of samples combined at the outset to a single batch. Given these three parameters we can calculate the expected number of total tests, *N*_*tests*_, for both of our methods, and thereby find the optimal batch-size, *b*, given *N* and *p*.

We show here that both approaches are roughly comparable to the absolute bound on test-efficiency, given by Shannon’s source code theorem. Furthermore, we compare these two approaches to matrix methods that attempt to exploit positional information on a 2D lattice in order to further increase efficiency. We find that without further optimization, matrix methods do not out-perform the simpler pooling methods presented here.

Finally, as a practical guideline for laboratories, we provide a table of optimal batch sizes for different ranges of *p*. In this practical solution, we round optimal batch sizes to multiples of 8, to fit the 8-raw based molecular tools common in most laboratories.

## Results

### Repeated Pooling

In the first method we present, we allow for repeated re-pooling of samples after each round of tests. While such a procedure in principle introduces additional parameters for the batch-sizes at each round of testing, we developed a simple, partially heuristic method based on a straightforward information-theoretic principle. The principle is that the most efficient binary test (one that, on average, excludes a maximal amount of possible scenarios) is one that has a 50% chance of giving either result (in our case – positive or negative for SARS-CoV-2). This is essentially a restatement of Shannon’s source coding theorem^6^, but it is also well-intuited by grade-school children through the games “Twenty Questions” and “Guess Who?”.

Our method is as follows: First, given the expected frequency of positive samples, *p*, calculate the initial batch-size, *b*, that yields as close to 50% positive rate. In all subsequent rounds of testing, simply divide any positive batches in half and repeat, essentially performing a binary tree search algorithm. The probability that an entire batch is negative is the product of the probabilities that each sample is negative. Thus, given *p*, the probability that an entire batch of size *b* is negative is *(1 - p)*^*b*^, and we want to choose *b* so that this number equals *0*.*5*. For example, suppose the frequency of positive samples is *p*=0.*02*, i.e. 1 in 50 samples is positive. It turns out that a batch of 34 samples has approximately a 50-50 chance of being entirely negative or having at least one positive, i.e. *(1 - p)*^*34*^ *≈ 0*.*5*. Therefore, we can write the desired initial batch-size *b*, as a function of *p*, as:

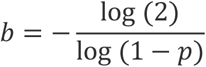

By fixing the batch size in this way it turns out that we also guarantee that with high probability there is only a single positive sample in each positive batch (Supplementary Figure). In the Appendix we derive the following estimate for the expected number of tests that need to be performed in this method, *Ntests* :

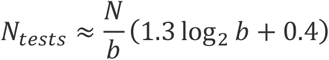

In our numerical simulations, as well as in our recommended protocols, we round the batch size, *b*, down to the nearest power of 2. For example, for *p*=0.*01*, the optimal initial batch size given by the expression above is 69 so we round it down to *b = 2*^*6*^ *= 64*. We find for these values of *p* and *b* that the average number of tests required to check *N* samples is about *N/8*. Meanwhile for *p*=0.*001*, the optimal initial batch size is *692*, which we round down to *b = 2*^*9*^ *= 512* in practice, and find that the average number of tests required to check *N* samples is about *N/50* (Figure 1). Figure 1 shows that our simulations with batch sizes rounded down to the nearest power of 2, are predicted very well by the two analytical expressions above (i.e. without rounding *b*).

**Figure 1.**
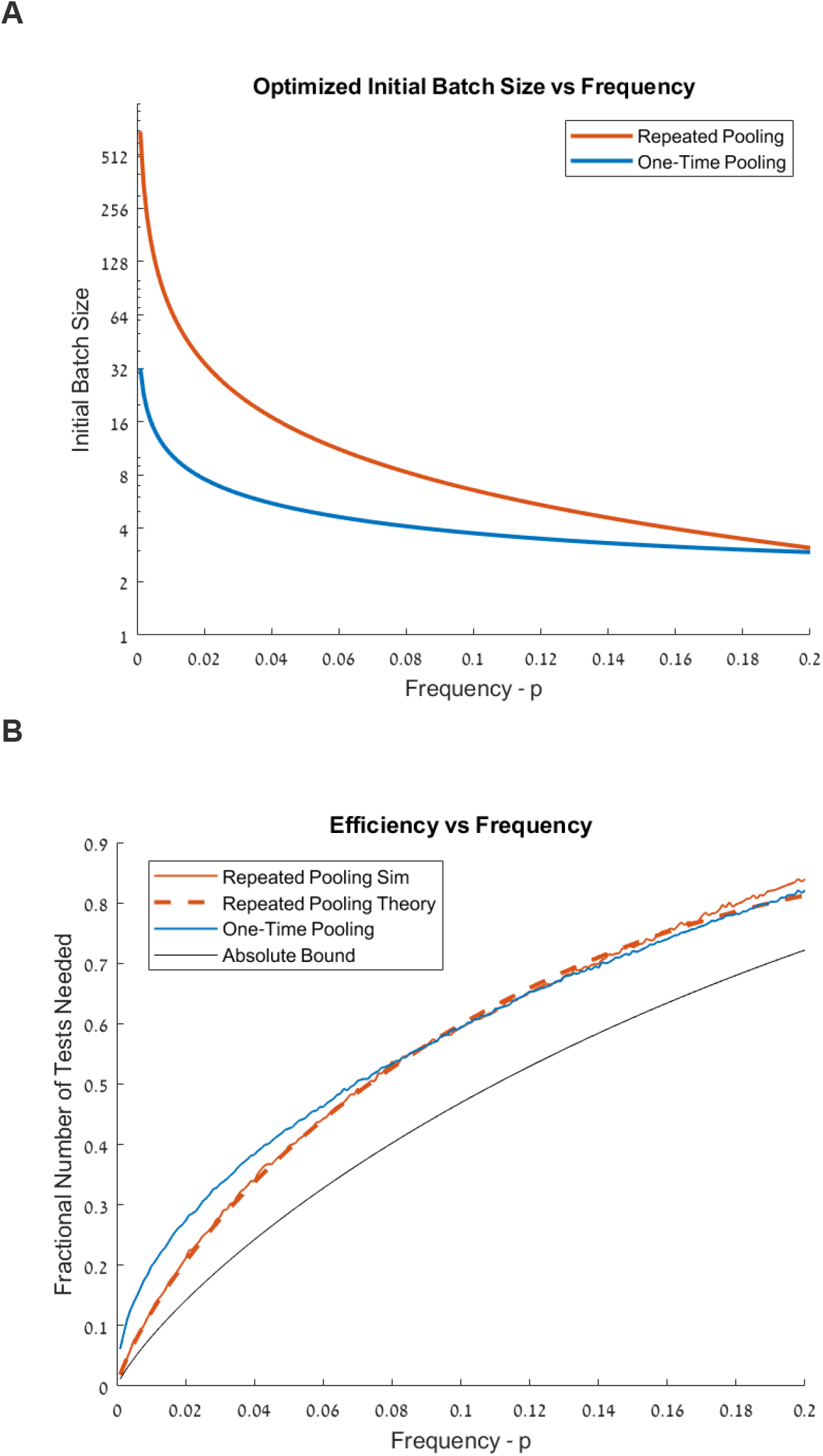
Optimal Batch Sizes and Expected Reduction in Tests Needed. **(A)** Optimal batch-size vs frequency of positive samples, with y-axis in log2 scale. Note than for in practice, for one-time pooling we round batch-sizes to the nearest whole number, and for repeated pooling we round batches down to the nearest power of 2. **(B)** Expected number of tests needed relative to the naïve approach of testing all samples, computed by repeatedly simulating large numbers of samples., Black line shows absolute minimum number of tests needed, given by the base-2 Entropy. *Blue:* Method of one-time pooling. *Red:* Method of repeated pooling. Dashed red line shows analytical calculation of expected number of tests via repeated pooling.

Technical limitations will clearly limit the maximal batch size. However, as we discuss further below, our simulations reveal that even suboptimal batch sizes are extremely efficient, especially for small values of *p*. For example, if the largest batch size is limited to 64, the number of tests required to test *N* samples with *p*=0.*001* is about *N/35*.

### One-time Pooling

Repeated pooling adds complexity to the protocol and may increase the likelihood of human error. We therefore propose a simpler method, proposed as early as 1943^3^: one-time pooling into batches, after which all samples in any positive batch are to be tested one-by-one. Under this method, very large batch sizes will no longer be as efficient, and we must calculate the optimal batch-size, *b*, specific to this method.

Given *b* and *p*, the expression for the expected number of tests under the one-time pooling method can be calculated as follows: First *N/b* tests must be performed. Next, how many of the total *N* samples will need to be tested individually? The expected fraction of total samples that must be tested individually is the same as the fraction of batches that is expected to be positive, because the batches are of equal size. The fraction of batches that is expected to be positive is simply the probability that any given batch will be positive: (*1 - (1 - p)*^*b*^). Thus, the total expected number of tests is:

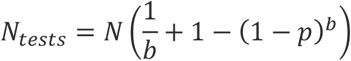

Consider the example in the introduction in which we have *N=1000* samples, *p*=0.*001*, and the batch size is *b=100*. Then we initially perform *N/b = 10* tests, and the probability that any entire batch is negative is *(1 - p)*^*b*^ *≈ 0.9*, so that the fraction of batches expected to be positive is *1/10*. We therefore perform an additional *N*/10 = 100 tests, for a total of *N(1/10 0 + 1/10) = ^11N^*/_100_ = 110.

The above expression can be optimized numerically to find the optimal *b* given p. We find for example, that for *p*=0.*01*, the optimal initial batch size is *b=*10, and the average number of tests required to check *N* samples is about *N/5*. Meanwhile for *p*=0.*001*, the optimal initial batch size is *b=32* and the average number of tests required to check *N* samples is about *N/16*. (Figure 1 and Table 1)

**Table 1.** One-Time Pooling

| Range of Ratios | Range of $p$ | Optimal Batch Size | Fraction of Tests Needed |
| --- | --- | --- | --- |
| <1:5 | $0.04 < p < 0.2$ | 4 | 0.40 - 0.84 |
| <1:25 | $0.008 < p < 0.04$ | 8 | 0.19 - 0.40 |
| <1:125 | $0.003 < p < 0.008$ | 16 | 0.11 - 0.18 |
| <1:333 | $0.001 < p < 0.003$ | 24 | 0.07 - 0.11 |
| <1:1000 | $0.0005 < p < 0.001$ | 32 | 0.05 - 0.06 |
| <1:2000 | $p < 0.0005$ | 64 | < 0.05 |

### Lower Bound on the Fractional Number of Tests

Shannon’s source coding theorem states that *N* independently and identically distributed random variables, each with entropy *H*, cannot be compressed into less than *NH* bits without loss of information^6^. The full series of binary diagnostic tests performed in our protocols can be thought of as an attempted compression of the *N* total samples. In our setting each sample is a Bernoulli(*p)* random variable and thus has entropy:

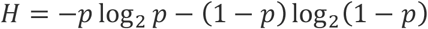

This therefore serves as the absolute lower bound for the fractional number of tests necessary, as a function of *p*. As shown in Figure 1, both of our proposed pooling strategies are roughly comparable to this lower bound.

### Robustness to Uncertainty

We have derived these two algorithms assuming perfect knowledge of the probability that any given sample is positive, *p*. It is important to check how robust our algorithms are to uncertainty. Therefore, we address two different kinds of uncertainty that need to be taken into account in everyday laboratory work.

First of all, even if the underlying probability of positive samples is known, the actual number of positive samples in a total of *N* samples will have a standard deviation that scales as 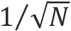. This intrinsic uncertainty will be significant for small *N*. To check robustness to intrinsic uncertainty in the actual frequency of positive samples, we simulate both algorithms for *N=128*. We find that this introduces a standard deviation of about 10 tests, and that this variability is in practice roughly independent of *p* and comparable between both methods, though slightly lower in the method of one-time pooling (Fig 2).

**Figure 2:**
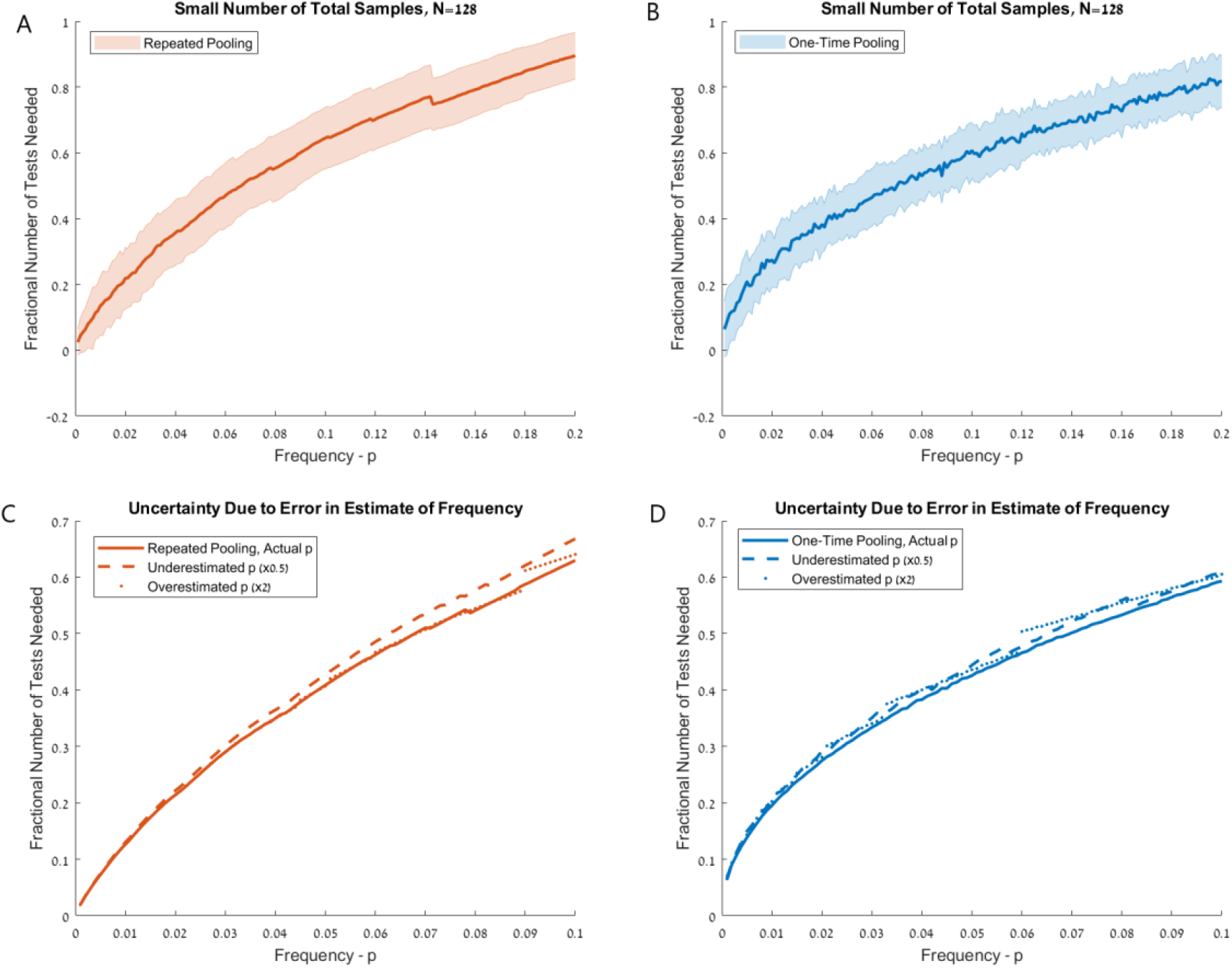
Robustness to Uncertainty. **(A)** Fractional number of tests needed relative to naïve approach, for repeated pooling method. Shaded area displays standard deviation over repeated trials with known frequency *p*, where variability is due to small number of total samples, *N=128*. **(B)** Same as (A), for one-time pooling. **(C)** Fractional number of tests via repeated pooling, with imprecise estimate of *p*. Dashed line displays results for *p* underestimated by a factor of two, and dotted line displays results for *p* overestimated by a factor of two. **(D)** Same as (C), for one-time pooling

An additional source of uncertainty is imperfect knowledge of the true probability, *p*. We find misestimating *p* by a factor of 2 in either direction has only small impact on the average number of tests needed by both of our methods (Fig 2). Thus, both algorithms are robust to the kinds of uncertainty expected in real laboratory settings.

Given this robustness, we propose simplified variations of the two methods, intended to overcome limited knowledge of *p*, and also minimize the complexity of the protocols. We propose that in practice, laboratories choose only a small number of possible initial batch sizes and identify the threshold value of *p* that separates between each initial batch size from the Tables below. For example, for the method of one-time pooling, a protocol that chooses between batch sizes of *b=16, b=8* or *b=4*, with thresholds of *p*=0.*008* and *p*=0.*04*, performs very well (Fig 3).

**Figure 3.**
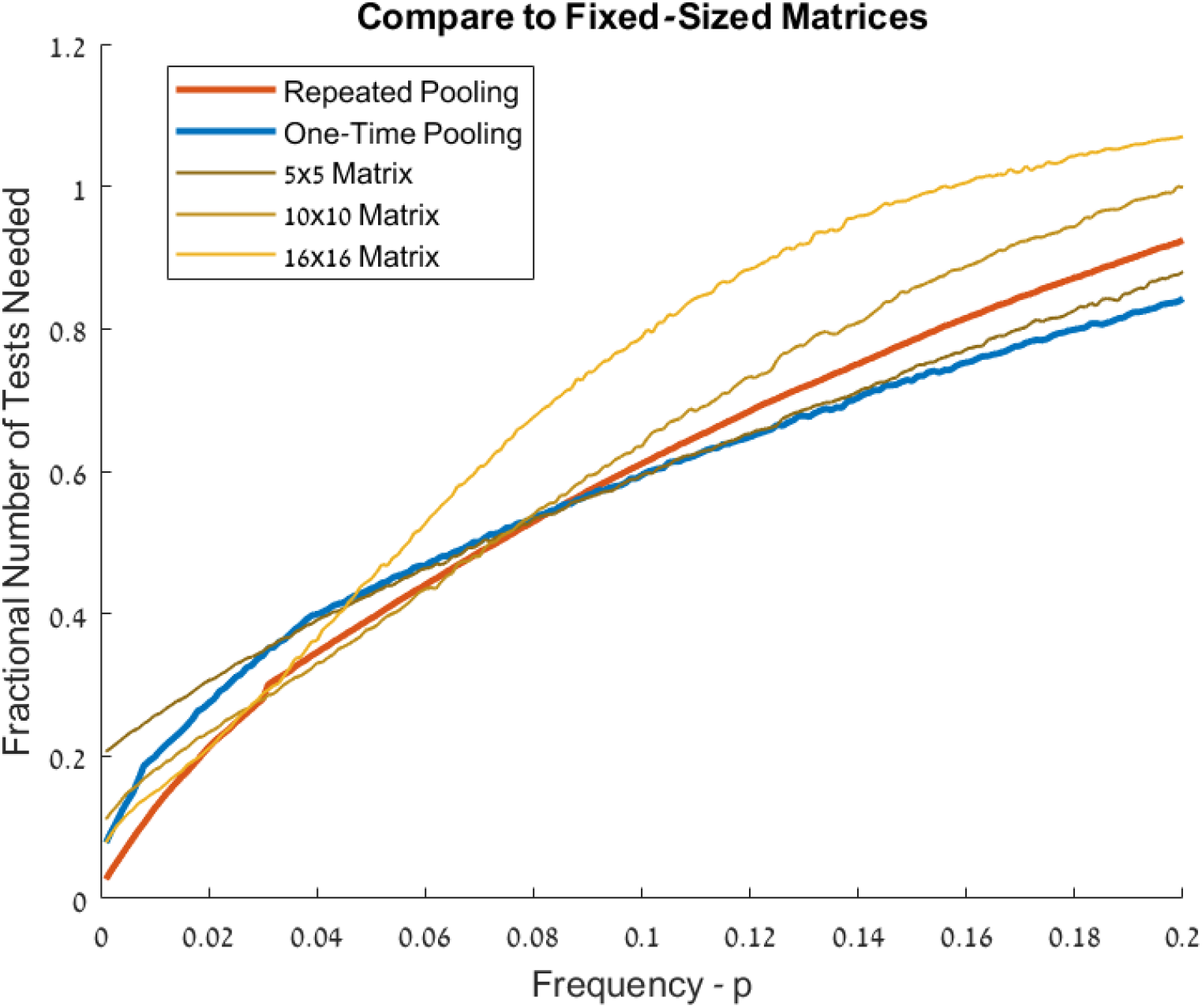
Comparison to Fixed-Size Matrix Method in Realistic Setting. We apply both methods presented here as they would be employed in a realistic setting with incomplete knowledge of *p*, and limited capacity for complex protocols. For one-time pooling we use three possible batch sizes: 4, 8 and 16. For repeated pooling we use four batch sizes: 8, 16, 32 and 64, with ranges of *p* assigned according to Tables 1 and 2, respectively. We compare the results to the matrix method, which uses 2D positional information, as described in the main text. No single method dominates all others. Note, however, that repeated pooling and one-time pooling can be easily combined into a single protocol with a single threshold value for *p*.

### Comparison to Matrix-based Methods

As an extension to pooling approaches, a number of matrix-based methods have been proposed that aim to exploit positional information on a 2D lattice of samples in order to further improve testing efficiency. The simplest of such approaches is as follows: assume samples are arranged on an *n*-x-*n* square grid, first pool each row as a single batch and perform *n* tests. If all *n* tests are negative stop, just as in the two methods proposed here. If any of the rows are positive, tests all *n* columns, and finally test all grid locations that are part of both positive row and positive column^9^. We find that fixed-size matrix methods do not out-perform the two methods presented here, even under realistic conditions of limited knowledge of *p* (Fig 3).

## Discussion

Efficient and rapid detection of SARS-CoV-2 is a crucial part of the global effort against the coronavirus pandemic. The use of RT-qPCR on a sample by sample basis is reportedly pushing the limits of available laboratory resources, such as chemical reagents. Here we present two pooling strategies that offer to dramatically reduce the use of such resources, as well as time and labor. In regions and testing conditions in which positive tests are very rare (*p*<0.05), a strategy of repeated pooling can be extremely efficient by first selecting an initial batch size that yields probability 0.5 of being entirely negative, and then proceeding by positive batches in half at each stage. As mentioned above, when positive samples are exceedingly rare this strategy in principle calls for very large batch sizes, well into the hundreds and even thousands. Such large batches are unfeasible with existing protocols. However, as we have shown, the strategy of repeated pooling is highly efficient in settings of exceedingly rare positives even when batch size is constrained to a pragmatic limit such as 64. Nevertheless, the process of repeatedly splitting pools into two may be challenging for many laboratories to implement in practice, and it loses marginal efficiency as the frequency of positive tests increases. We therefore show that a simpler protocol of one-time pooling, with optimized initial batch sizes is very efficient for all *p* up to about 0.2. One-time pooling is efficient even when the size of possible initial batch sizes is technically limited, for example to 16, either in order to simplify laboratory protocol or because knowledge of *p* is lacking. We show that both methods compare favorably to fixed-size matrix methods, that attempt to exploit 2D positional information from samples arranged on a grid. We note, however, that matrix methods can in principle also be optimized for given *p*, or by using complex pooling strategies^10^. Optimized matrix methods may prove more efficient than the two straight-forward methods presented here.

It is important to note that technical limitations may limit the maximal batch size. This is because the process of pooling multiple patient samples into one tube inevitably causes dilution of the RNA of each individual sample. While it was shown empirically that 64 samples could be pooled together to a combined sample that contains enough RNA copies for detection^2^, further empirical work should be conducted in order to determine the maximal pool possible. For very large pools, improvement could be achieved with a minor change in the existing protocol (e.g. extracting higher concentrations of RNA content perhaps at the expense of some background reagents). Nevertheless, we chose here to show the theoretical optimal batch size, even if its feasibility is still somewhat questionable. To keep our methods implementable immediately, we calculate their performance also for a constrained batch size and present these in the practical tables and protocols. Note, as we write in our protocols, that even moderate batch sizes require an appropriate adjustment of the cycle-threshold for detection.

We hope this study will assist to increase the number of tests thus improving local governments’ and agencies’ ability to monitor and prevent the spread of COVID-19.

## Protocols and Tables

- First, the laboratory should select the pooling method. As mentioned, one-time pooling should be favored for its simplicity and smaller optimal batch sizes, except in cases where *p* is very low (p<0.05) and the laboratory expertise allows multiple pooling steps without substantial risk of errors.
- Next, define the maximal batch size from the appropriate table below, based on the laboratory’s technical considerations.
- At the start of each round of tests, estimate the value of p. This value should be estimated and updated on a regular (e.g. daily) basis, according to the empirical results received in a specific lab/region/population. It is simply calculated as follows: *p* = (number of positive samples)/ (number of all tested samples).
- Next, identify the initial batch size from the tables below, by finding the appropriate range of *p*. Each laboratory should ignore ranges (i.e. rows) associated with batch sizes larger than the laboratory’s chosen maximum, and in cases where *p* is estimated in those ranges simply choose the maximal batch-size. For example, a protocol of one-time pooling with maximal batch size of *16* would set an initial batch size of *4* for *p>0.04*, a batch size of *8* for *p* between 0.008 and 0.04, and a batch size of 16 for all *p<0.008*.
- Finally, before performing RT-qPCR, increase the cycle-threshold by *log*_*2*_ *b* cycles. This is because pooling, if uncompensated, will dilute the presence of RNA/cDNA by a factor of *b*, which is expected to delay threshold-crossing. For example, if viral RNA is diluted by a factor of *8* from the original concentration in a sample then it will take 3 cycles of doubling in order to reach the original concentration. This should be taken into consideration when selecting maximal batch size.

### One-Time Pooling

According to the estimated value of *p*, use Table 1 to decide on the initial batch size. Process all batches/pools through PCR. All samples from batches that were shown to be negative could be all regarded as negatives, and all samples in batches that were shown to be positive should be further processed individually. Laboratories should restrict the number of possible initial batch sizes by simply ignoring ranges of smaller *p*, i.e. larger batch sizes. Table 1 summarizes also the expected fraction of tests needed relative to the naïve approach of testing all samples. This is important for the planning and evaluating the expected gain of the selected method beforehand.

### Repeated Pooling

According to the estimated value of *p*, use Table 2 to decide on the optimal initial batch size. Process all batches/pools through PCR. All samples from batches that were shown to be negative could be all regarded as negatives. Batches that were shown to be positive should be further divided into equal sizes (for odd numbers just chose arbitrary one batch to be bigger than the other by 1 sample), until all positive samples are isolated. In practice, positive batches of size 4 or smaller should not be divided, but simply tested individually.

**Table 2.** Repeated Pooling

| Range of Ratios | Range of $p$ | Optimal Batch Size | Fraction of Tests Needed |
| --- | --- | --- | --- |
| <1:20 | $0.03 < p < 0.05$ | 8 | ~0.4 |
| <1:33 | $0.02 < p < 0.03$ | 16 | ~0.3 |
| <1:50 | $0.01 < p < 0.02$ | 32 | ~0.2 |
| <1:100 | $0.005 < p < 0.01$ | 64 | ~0.13 |
| <1:200 | $0.0025 < p < 0.005$ | 128 | ~0.08 |
| <1:400 | $0.001 < p < 0.0025$ | 256 | ~0.04 |
| <1:1000 | $0.0005 < p < 0.001$ | 512 | ~0.02 |
| <1:2000 | $p < 0.0005$ | 1024 | ~0.01 |

Use the expected fraction of tests needed relative to naïve approach for evaluation of the expected gain of the method beforehand.

## Code and Data

Code and simulation data is available at https://github.com/ilandau/sample-pooling-covid19.

## Data Availability

All data and code is available here at the link provided.

https://github.com/ilandau/sample-pooling-covid19

## Acknowledgements

We thank Prof. Eran Meshorer for very helpful comments.

## Notes

### Competing Interest Statement

The authors have declared no competing interest.

### Funding Statement

No external funding was received.

## Bibliography

1. Hossain et al “A Massively Parallel COVID-19 Diagnostic Assay for Simultaneous Testing of 19200 Patient Samples”. (2020 preprint) https://docs.google.com/document/d/1kP2w_uTMSep2UxTCOnUhh1TMCjWvHEY0sUUpkJHPYV4/edit

2. Yedin, I. et al.. “Evaluation of COVID-19 RT-qPCR test in multi-sample pools”. medRxiv (2020) https://doi.org/10.1101/2020.03.26.20039438

3. Dorfman, R. “The detection of defective members of large populations”. In: The Annals of Mathematical Statistics 14.4 (1943)

4. Gaidet, N. et al.. Saving resources: Avian influenza surveillance using pooled swab samples and reduced reaction volumes in real-time RT-PCR. Artic. J. Virol. methods 186, 119–125 (2012).

5. CDC 2019-Novel Coronavirus (2019-nCoV) Real-Time RT-PCR Diagnostic Panel. <https://www.cdc.gov/coronavirus/2019-ncov/lab/index.html>

6. Cover, TM (2006). “Chapter 5: Data Compression”. Elements of Information Theory. John Wiley & Sons. ISBN 0-471-24195-4

7. https://covidtracking.com

8. https://ourworldindata.org/coronavirus-testing-source-data

9. Sinnott-Armstrong, N et al. “Evaluation of Group Testing for SARS-CoV-2 RNA”. medRxiv (2020) https://doi.org/10.1101/2020.03.27.20043968

10. Erlich, Y et al.. “DNA Sudoku - Harnessing high throughput sequencing for multiplexed specimen analysis”. In: Genome Research 19.7 (2009), pp. 1243–1253

11. https://www.newyorker.com/news/news-desk/why-widespread-coronavirus-testing-isnt-coming-anytime-soon

12. https://www.thetimes.co.uk/article/global-shortage-of-coronavirus-testing-kits-threatens-expansion-plan-chglmtm93

